# Clinical Utility of the Molecular Microscope Diagnostic System in a Real-World Transplant Cohort: Moving Towards a New Paradigm

**DOI:** 10.1101/2024.06.24.24309444

**Authors:** Andrea Fernandez Valledor, Cathrine M. Moeller, Gal Rubinstein, Salwa Rahman, Daniel Oren, Julia Baranowska, Changhee Lee, Ruben Salazar, Carolyn Hennecken, Afsana Rahman, Boaz Elad, Dor Lotan, Ersilia M. DeFilippis, Adil Yunis, Justin Fried, Jayant Raihkelkar, Kyung T. Oh, David Bae, Edward Lin, Sun Hi. Lee, Matthew Regan, Melana Yuzelpolskaya, Paolo Colombo, David T. Majure, Farhana Latif, Kevin D. Clerkin, Gabriel T. Sayer, Nir Uriel

## Abstract

**Objectives:** To evaluate the clinical implications of adjunctive molecular gene expression analysis (MMDx) of biopsy specimens in heart transplant (HT) recipients with suspected rejection.

**Introduction:** Histopathological evaluation remains the standard method for rejection diagnosis in HT. However, the wide interobserver variability combined with a relatively common incidence of “biopsy-negative” rejection has raised concerns about the likelihood of false-negative results. MMDx, which uses gene expression to detect early signs of rejection, is a promising test to further refine the assessment of HT rejection.

**Methods:** Single-center prospective study of 418 consecutive *for-cause* endomyocardial biopsies performed between November 2022 and May 2024. Each biopsy was graded based on histology and assessed for rejection patterns using MMDx. MMDx results were deemed positive if borderline or definitive rejection was present. The impact of MMDx results on clinical management was evaluated. Primary outcomes were 1-year survival and graft dysfunction following MMDx-guided clinical management. Secondary outcomes included changes in donor-specific antibodies, MMDx gene transcripts, and donor-derived cell-free DNA (dd-cfDNA) levels.

**Results:** We analyzed 418 molecular samples from 237 unique patients. Histology identified rejection in 32 cases (7.7%), while MMDx identified rejection in 95 cases (22.7%). Notably, in 79 of the 95 cases where MMDx identified rejection, histology results were negative, with the majority of these cases being antibody-mediated rejection (62.1%). Samples with rejection on MMDx were more likely to show a combined elevation of dd-cfDNA and peripheral blood gene expression profiling than those with borderline or negative MMDx results (36.7% vs 28.0% vs 10.3%; p<0.001). MMDx results led to the implementation of specific antirejection protocols or changes in immunosuppression in 20.4% of cases, and in 73.4% of cases where histology was negative and MMDx showed rejection. 1-year survival was better in the positive MMDx group where clinical management was guided by MMDx results (87.0% vs 78.6%; log rank p=0.0017).

**Conclusions:** In our cohort, MMDx results more frequently indicated rejection than histology, often leading to the initiation of antirejection treatment. Intervention guided by positive MMDx results was associated with improved outcomes.

**Graphical abstract:** 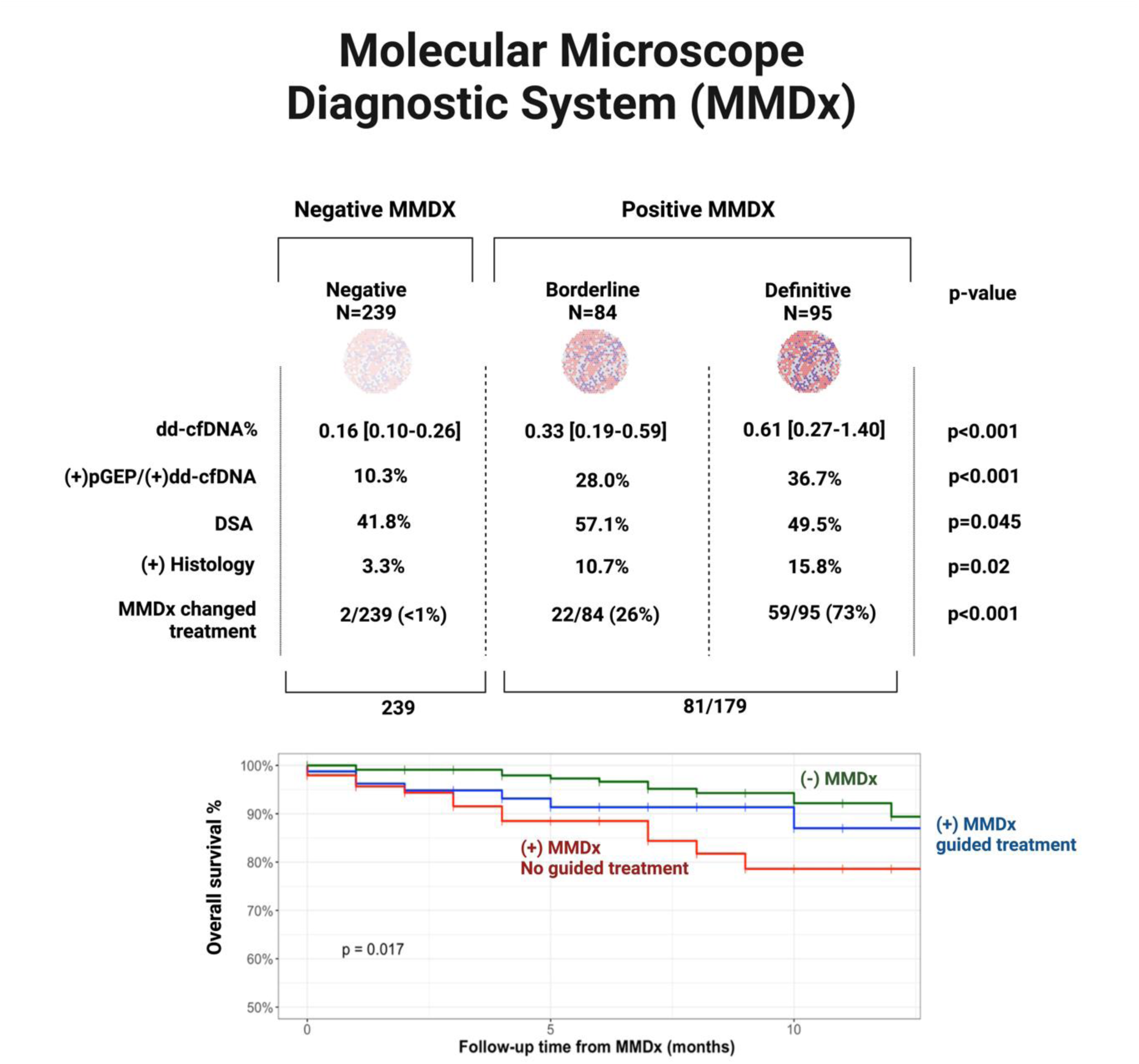

## INTRODUCTION

Heart transplantation (HT) remains the preferred therapy for patients with advanced heart failure who do not exhibit features of myocardial recovery and lack contraindications (1). Despite refinements in immunosuppressive schemes, allograft rejection remains a compelling cause of morbidity in the initial stages, and one of the leading determinants of long-term graft survival (1–4). Although the gold standard for rejection diagnosis is the histological assessment of the endomyocardial biopsy (EMB), its reliability is hindered by the reliance on expertise in pattern recognition, the lack of consensus on complement-independent antibody-mediated rejection pathways, and the high prevalence of biopsy-negative rejection (5–8).

Over the past few years, we have witnessed a rise in the adoption of novel rejection surveillance tools, such as donor-derived cell-free DNA (dd-cfDNA) and the Molecular Microscope Diagnostic System (MMDx, One Lambda, CA) (6,9–13). These advancements have enabled the early detection of rejection even before histological changes become apparent (14). As a result, physicians increasingly encounter situations with discrepant results in which the role of the clinician becomes more indispensable than ever, as multiple variables need to be incorporated into therapeutic decision-making. The molecular diagnosis provided by MMDx offers a probabilistic assessment of rejection independent of histological findings, enhancing diagnostic accuracy with high precision (6). Supervised molecular classifiers have demonstrated superior predictive ability for molecular rejection, with areas under the curve > 0.87, compared to histologic rejection < 0.78 (11). Previous research has examined the correlation between MMDx and histology as well as the association between dd-cfDNA and MMDx-rejection related transcripts (15–18). However, there is still limited understanding of how integrating MMDx into clinical decision-making processes can impact patient care. Therefore, we aimed to assess the clinical utility of MMDx analysis in a cohort of heart transplant recipients undergoing EMB for clinical indication.

## METHODS

### Study design and population

Consecutive HT recipients that underwent EMB due to suspected rejection between November 2022 and May 2024 at our institution were prospectively included. This study was approved by the Institutional Review Board with a waiver of informed consent. Causes for biopsy included: 1) clinical symptoms compatible with rejection (e.g. heart failure symptoms or chest pain); 2) de-novo allograft dysfunction [defined as left ventricular ejection fraction (LVEF) less than 50%, or a decline in LVEF >10% from the previous echocardiography (TTE); 3) elevated dd-cfDNA levels (≥0.12%, Allosure, CareDX); 4) de novo or rising donor specific antibodies (DSA), 5) elevated biomarkers (NT-proBNP and/or high-sensitivity troponin T), and/or 6) follow-up on recently treated rejection. EMB was performed in the standard fashion with 5-6 samples obtained. Three tissue samples were sent for histological evaluation and immunohistochemistry, 1 sample was sent for immunofluorescence and between one to two samples were sent for molecular analysis (MMDx).

### Data collection

Baseline characteristics, transplant data information, current immunosuppressive regimen, echocardiography assessment, presence and type of DSA, cardiac biomarkers and hemodynamics at the time of biopsy, were systematically collected for each sample. This also included paired dd-cfDNA levels and peripheral blood gene-expression profiling (pGEP). Viral polymerase chain reactions for Ebstein-Barr virus (EBV), BK and cytomegalovirus (CMV) were also systematically checked at the time of pGEP/ dd-cfDNA screening.

### Non-invasive monitoring

Since 2019, our center has used a non-invasive rejection surveillance protocol in which dd-cfDNA and pGEP testing are standard of care, commencing at the 6th week for dd-cfDNA and at the 8th week for pGEP. These tests are conducted monthly during the first-year post-transplant and then shift to a quarterly schedule until the 3rd year post-transplant. After the first two years, additional dd-cfDNA and pGEP samples are collected as clinically indicated. At our institution, positive dd-cfDNA levels were defined as ≥0.12%. For the purposes of this study, we report results for the ≥0.12% threshold as well as the ≥0.20% threshold, which has been used to define rejection in other studies. We used two thresholds for pGEP positivity: >30 within the first 5 months after transplant and >34 for ≥6 months post-transplant. Only dd-cfDNA levels and pGEP results obtained within 31 days of biopsy and those from single organ recipients were included in the analysis.

### Donor specific antibodies (DSA)

Our protocol includes the evaluation of DSA at specific intervals: weeks 1, 4, and months 3, 6, 9, 12, followed by every six months after the first year using the Luminex assay. The threshold for antibody detection is set at medium fluorescence intensity (MFI) > 1000. DSA were classified as class I or II antibodies and further categorized based on MFI levels as follows: low (<4000), moderate (4000-10000), high (10000-20000) and very high (>20000). We also routinely screened patient for major histocompatibility complex class I chain-related gene A (MIC-A) antibodies.

### Hemodynamics

At the time of EMB, right heart catheterization was routinely performed, with standard assessment of hemodynamics and using the assumed Fick method to calculate cardiac output. Abnormal hemodynamics were defined as a cardiac index <2.2 l/min*m^2^, mean pulmonary pressures > 24 mmHg, pulmonary capillary wedge pressure >15 mmHg or right atrial pressure >8 mmHg.

### Histological assessment

Histological evaluation was performed by three pathologists who were blinded to clinical information. All biopsies were routinely evaluated for histopathologic evidence of acute cellular rejection (ACR), and antibody-mediated rejection (pAMR). All biopsies were screened for complement deposition (C4d) and intravascular monocyte presence (CD68). Biopsies were graded based on the criteria outlined in the International Society of Heart and Lung Transplant (ISHLT) 2004 guidelines (19) for acute cellular rejection and the revised 2013 guidelines for AMR (20–21). Histological rejection was identified as positive if ISHLT grading>1R/1A rejection was present and/or pAMR > 0. In cases where positive results were followed by treatment, a repeat biopsy was typically conducted within two weeks for ACR and within four to eight weeks after antibody-mediated rejection treatment.

### Molecular Microscope Diagnostic System (MMDx)

MMDx is a central diagnostic system that uses microarrays to measure mRNA levels in transplants biopsies. Archetype scores for normal (NRI), T-cell mediated rejection (TCMR), antibody-mediated rejection (ABMR) and injury that go from 0 to 1 are given for each biopsy. The models translate these archetypes into rejection probabilities. Generally, a definitive diagnosis is established when the probability exceeds 0.6. Intermediate cases in which the inflammation does not reach the extent to be considered as definite rejection are defined as borderline rejection. The MMDx report provides additional molecular data including pathogenesis-based transcript set scores and transcript expression scores relating to the different rejection subtypes and parenchymal injury. For each score, a normal limit is given, defined as the 95^th^ percentile score in the normal biopsies (6). “Rejection” or “definitive rejection” on MMDx was defined as the presence of TCMR, ABMR, or mixed rejection. “Positive” MMDx results included borderline rejection along with definitive rejection due to the similarities of borderline cases to those with definitive rejection. Cases classified as normal (NRI) or with parenchymal injury were considered negative MMDx.

### Clinical decision making

The multidisciplinary care team integrated clinical presentation, histological and molecular results, dd-cfDNA levels and pGEP scores, biomarkers, graft function and hemodynamics, presence and trajectory of DSA, history of rejection episodes and past treatments to determine subsequent treatment plans. The final decision is not protocolized and is left to the discretion of clinicians. **Figure 1**.

**Figure 1:**
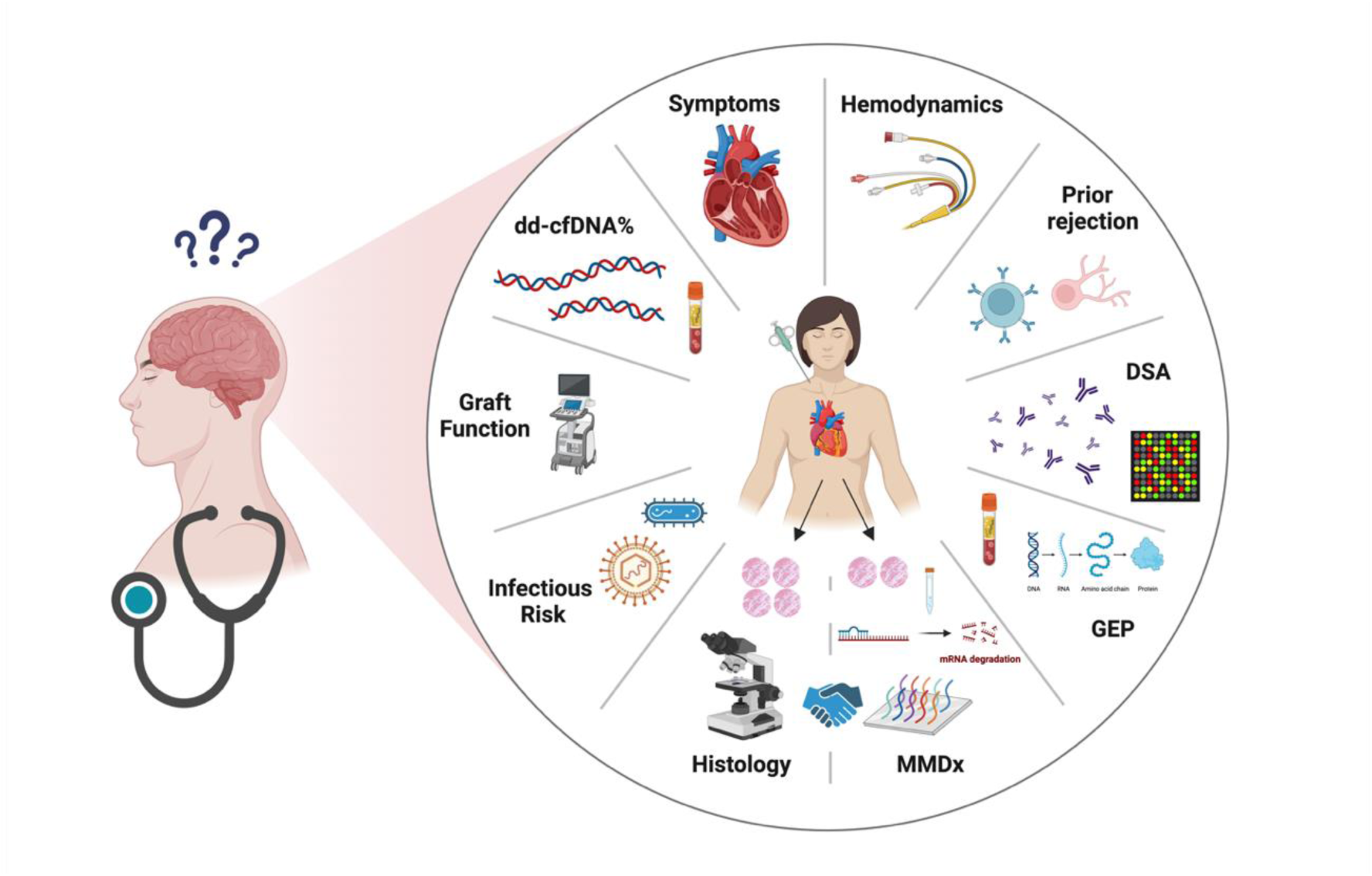
multifaceted rejection approach.

### Outcomes

We evaluated how the implementation of MMDx influenced clinical management and its impact on outcomes. Specifically, we compared 1-year survival rates and the incidence of graft dysfunction based on MMDx results and subsequent MMDx-guided interventions. Graft dysfunction was defined as a new occurrence of LVEF <50% or a decline in LVEF >10% during the follow-up period. Patients already exhibiting graft dysfunction at the time of MMDx were censored from this analysis. Secondary outcomes included changes in MFI levels of DSA, MMDx-transcripts, and dd-cfDNA in the subsequent screening following treatment adjustment based on MMDx results. Improvement in DSA was defined as a reduction in MFI category (e.g., from high to moderate) or a 25% reduction in MFI levels if the baseline MFI was >4000. We also monitored the development of de novo DSA if MFI was >2000. Additionally, we measured changes in kidney function, and infection rates within three months following the increase in immunosuppression.

### Data Analysis and Statistical Methods

Statistical analysis was performed using R Core Team (2020): A language and environment for statistical computing, R Foundation for Statistical Computing, Vienna, Austria version 4.1.3. Scatter and bar plots were created using Microsoft Office Excel. The Shapiro-Wilk test was performed on all continuous variables to determine normality. Gaussian distributed continuous variables were reported as means and standard deviations. Non-gaussian distributed variables were reported as median and interquartile range. Categorical variables were summarized as n (%) and compared using Fisher’s Exact Test or chi square test as appropriate for independent variables or McNemar’s test for paired variables. Parametric variables were compared using independent two-sample or paired sample t-test as appropriate. Non-parametric variables were compared using Kruskal-Wallis test for independent samples, and Friedman test for dependent samples. A p-value of 0.05 was considered statistically significant. Survival analysis was conducted using the Kaplan Meier method to identify overall survival and freedom from graft dysfunction at 1-year stratified based on the MMDx result and management. A log-rank test was employed to evaluate the difference between the survival curves.

## RESULTS

During the study period, 441 paired histology and MMDx EMB samples were identified. Of these, 23 were excluded as they were performed as part of our surveillance monitoring protocol. The final cohort consisted of 418 samples from 248 heart transplant recipients. **Figure 2**.

**Figure 2:**
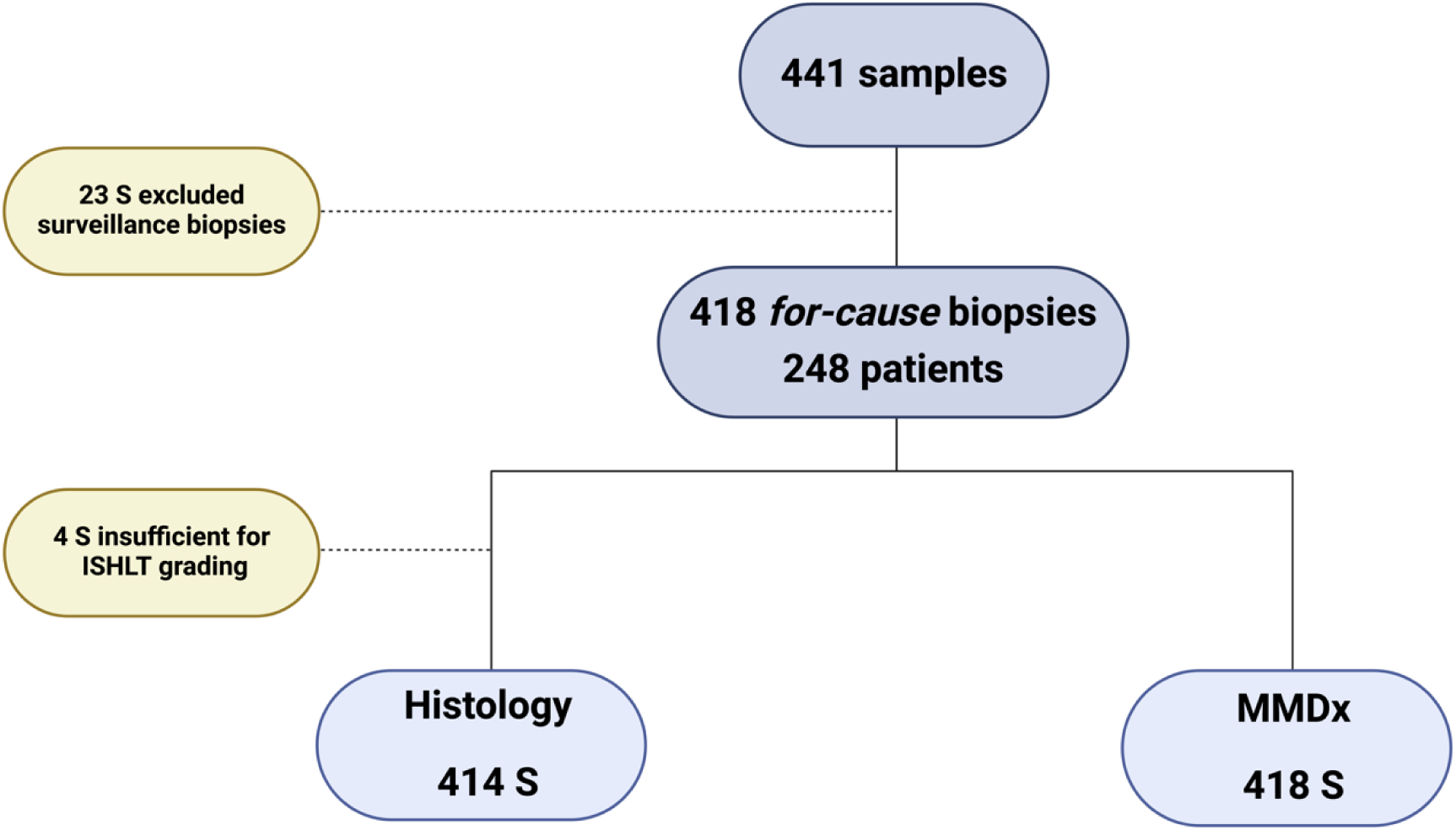
flow chart.

### Baseline Characteristics

Patient baseline characteristics are represented in **Table 1S.** The leading indication for biopsy was an elevated dd-cfDNA levels in 40.2% of the cases, followed by symptoms (21.1%), rising DSA level (13.4%), de-novo or worsening allograft dysfunction (12.2%), follow-up on recently treated rejection (11.2%) and elevated biomarkers (1.9%).

### Concordance and Discordance between Histology and MMDx

Concordance and discordance between histology and MMDx are shown in **Figure 3A**. Histological analysis detected rejection in 32 samples (7.7%), whereas MMDx identified rejection in 95 samples (22.7%). The overall concordance between histology and MMDx was 76.8% (S=318), primarily driven by negative concordance (73.2%; S=303). Of the 96 samples with discordant results (23.2%), 79 samples (82.3%) had negative histology, but rejection detected by MMDx. In this group, MMDx identified ABMR in 48 samples (60.7%), TCMR in 26 samples (32.9%), and mixed rejection in 5 samples (6.3%). **Figure 3A**. Of the 17 cases (16.0%) with positive histology and negative MMDx, 9 (53.0%) exhibited low-grade rejection on histology (2 cases of 1B rejection and 7 cases of pAMR=1i). **Table 2S.**

**Figure 3:**
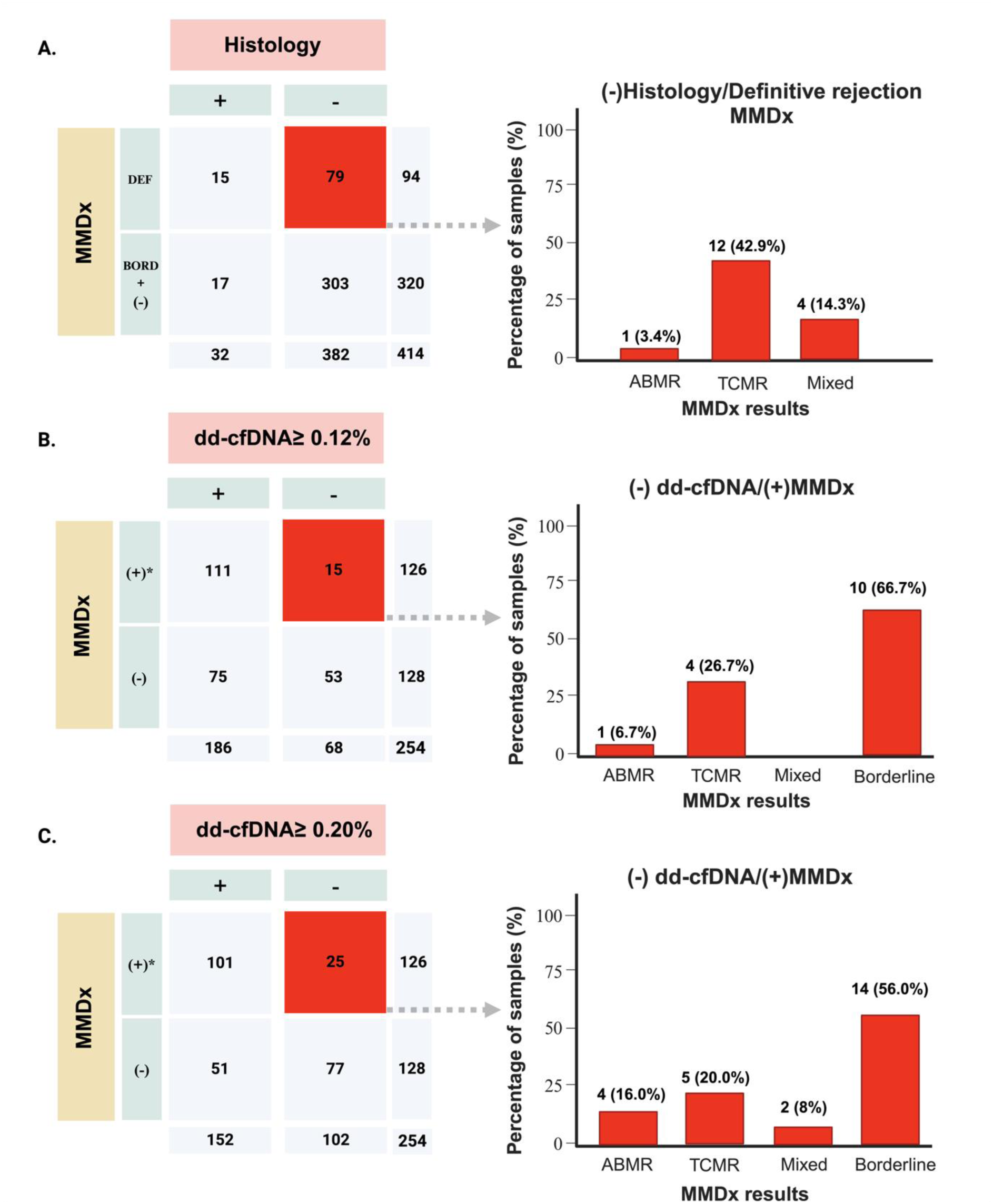
(**A**) illustrates the concordance between histology and MMDx results as well as between dd-cfDNA levels and MMDx results at both thresholds [≥0.12% (**B**) and ≥0.20% (**C**)]; “DEF” accounts for definitive rejection, which includes ABMR, TCMR, and mixed rejection cases, while “BORD” refers to borderline rejection. *(+) MMDx results include definitive and borderline rejection cases, whereas (-) MMDx includes cases of normal results and parenchymal injury.

A trend towards a higher prevalence of DSA was observed when both histology and MMDx indicated rejection (77.3%), compared to cases where only histology (64.7%) or only MMDx (44.3%) indicated rejection (p=0.07). However, the presence of moderate-level or greater DSA (MFI >4000) was more common when both techniques detected rejection compared to cases where only one method detected rejection (p<0.001). **Table 2S.**

### Concordance and Discordance between dd-cfDNA and MMDx

Concordance and discordance between dd-cfDNA levels at 0.12% and 0.20% thresholds with MMDx are depicted in **Figures 3B** and **3C** respectively. When comparing positive MMDx results to dd-cfDNA levels, the concordance was higher using the 0.20% threshold (70.1%) compared to the 0.12% threshold (64.6%). This improvement in concordance was mainly due to an increase in negative concordance at the higher threshold.

### MMDx results

Samples in which MMDx showed rejection were further out from transplant and had a higher prevalence of prior treated rejections than samples with borderline or negative MMDx results. **Table 1**. Positive dd-cfDNA results were observed in 92.6% and 83.8% of cases with rejection on MMDx at ≥0.12% and ≥0.20% thresholds, respectively. dd-cfDNA levels were also higher in cases of definitive rejection (0.61% [0.27-1.40]) compared to those with borderline (0.33% [0.19-0.59]) or negative MMDx results (0.20% [0.14-0.34]; p<0.001). **Figure 1S**. That contrast with histology, where dd-cfDNA levels did not significantly differ between rejection and no-rejection samples (0.44% [0.20-1.70] vs. 0.24% [0.11-0.52]; p=0.06).

**Table 1:**
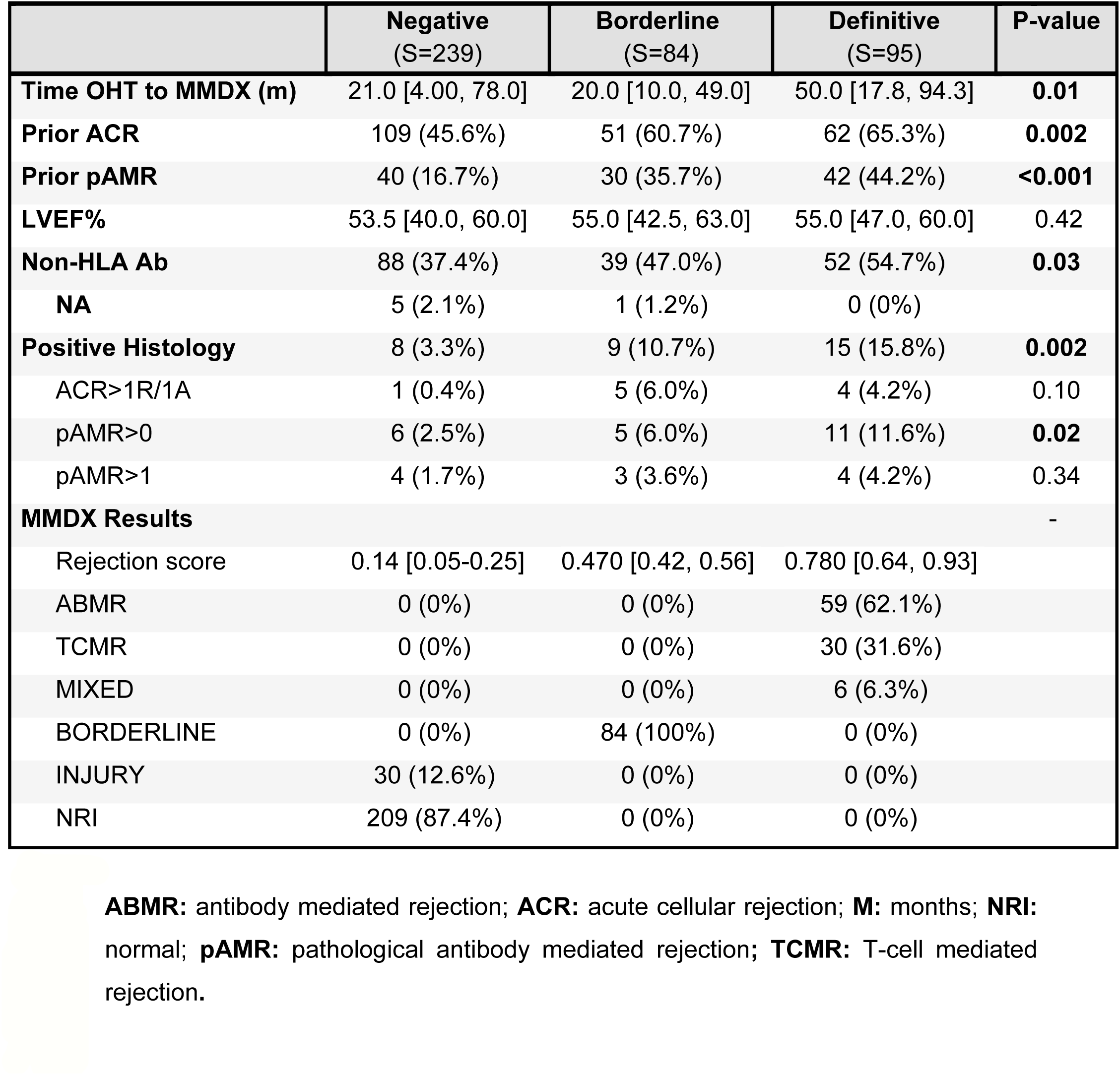
sample characteristics stratified by MMDx results.

When combined with pGEP, samples with rejection on MMDx had higher rates of combined (+)pGEP/(+)dd-cfDNA (36.7%) compared to samples with borderline (28.0%) or negative MMDx results (10.3%); p<0.001. Specifically, samples with (+)pGEP/(+)dd-cfDNA had three times higher rates of definitive rejection detected by MMDx (36.7% vs 10.3%; p<0.001). Furthermore, when compared to samples with (-)pGEP/(-)dd-cfDNA, the presence of (+)pGEP/(+)dd-cfDNA was associated with a fourfold increase in the rates of definitive rejection on MMDx (36.7% vs 8.3%; p<0.001). **Table 2**.

**Table 2:** non-invasive biomarkers and DSA stratified based on MMDx results.

|  | Negative<br>(S=239) | Borderline<br>(S=84) | Definitive<br>(S=95) | P-value |
| --- | --- | --- | --- | --- |
| **dd-cfDNA≥ 0.12%** | 75 (58.6%) | 48 (82.7%) | 63 (92.6%) | **<0.001** |
| **dd-cfDNA≥ 0.20%** | 51 (39.8%) | 44 (75.9%) | 57 (83.8%) | **<0.001** |
| NA | 111 (46.4%) | 26 (31.0%) | 27 (28.4%) |  |
| **dd-cfDNA levels [%]** | 0.16 [0.10, 0.26] | 0.33 [0.19, 0.59] | 0.61 [0.27, 1.40] | **<0.001** |
| **Time dd-cfDNA to EMB (d)** | 10.0 [6.00, 18.3] | 12.0 [8.0, 16.0] | 10.5 [6.00, 16.3] | 0.52 |
| **pGEP levels** | 31.0 [27.0, 34.0] | 33.0 [29.0, 35.0] | 33.0 [31.8, 36.0] | **<0.001** |
| **Positive pGEP** | 22 (20.0%) | 20 (39.2%) | 26 (43.3%) | **0.002** |
| NA | 129 (54.0%) | 33 (39.3%) | 35 (36.8%) |  |
| **Time pGEP to EMB (d)** | 10.0 [6.00, 15.8] | 11.0 [7.50, 16.0] | 11.5 [6.75, 16.0] | 0.41 |
| **HeartCare** |  |  |  | **<0.001** |
| (-)pGEP/(-)dd-cfDNA | 50 (46.7%) | 6 (12.0%) | 5 (8.3%) | **<0.001** |
| (-)pGEP/(+)dd-cfDNA | 35 (32.7%) | 24 (48.0%) | 29 (48.3%) | 0.07 |
| (+)pGEP/(-)dd-cfDNA | 13 (5.4%) | 6 (12.0%) | 4 (6.7%) | 0.51 |
| (+)pGEP/(+)dd-cfDNA | 11 (10.3%) | 14 (28.0%) | 22 (36.7%) | **<0.001** |
| NA | 129 (54.0%) | 33 (39.3%) | 35 (36.8%) |  |
| **DSA** | 100 (41.8%) | 48 (57.1%) | 47 (49.5%) | **0.045** |
| Class I | 39 (16.3%) | 22 (26.2%) | 23 (24.2%) | 0.09 |
| Class II | 87 (36.4%) | 44 (52.4%) | 39 (41.1%) | **0.049** |
| MFI>4000 | 48 (20.1%) | 29 (34.5%) | 27 (28.4%) | **0.02** |
| **Time DSA to EMB (d)** | 8.00 [2.00, 20.0] | 9.00 [0.750, 16.0] | 8.00 [3.00, 20.5] | 0.46 |

**D:** days; **dd-cfDNA:** donor-derived cell-free DNA; **DSA:** donor specific antibodies; **EMB:** endomyocardial biopsy; **pGEP:** peripheral gene-expression profiling; **MFI:** medium fluorescence intensity.

The prevalence of positive DSA and moderate strength or greater DSA was similar between cases of definitive rejection and borderline MMDx results (49.5% vs. 57.1%; p = 0.30 and 28.4% vs. 34.5%; p = 0.38, respectively). However, these prevalences were significantly higher compared to samples with negative MMDx (p=0.045 for DSA and p=0.02 for higher MFI); **Table 2**. The presence of de-novo graft dysfunction (p=0.82) or abnormal hemodynamics (p=0.34) at the time of biopsy was similar between the groups. **Table 3S**. Samples with rejection were more commonly from patients receiving belatacept (27.4% vs 10.7% vs 7.9%; p<0.001) or undergoing extracorporeal photopheresis (ECP) (7.4% vs 3.6% vs 0.8%; p=0.006) as part of their immunosuppressive regimen compared to samples with borderline or negative MMDx results. **Table 3S.**

### MMDx and clinical decision making

The addition of MMDx results altered clinical management in 20.3% of cases (84/414). **Figure 4**. In instances of negative histology and rejection on MMDx (N=79), immunosuppressive therapy was altered in 73.4% of cases (58/79). Specifically, 17 patients (29.3%) underwent acute ABMR protocols, which included bortezomib, plasmapheresis (PLEX), and intravenous immunoglobulin (IVIg) in 11 cases; thymoglobulin, PLEX, IVIg, and rituximab in 2 cases; or rituximab and IVIg in 4 cases. Sixteen patients (27.6%) received pulse steroids alone, and 25 patients (43.1%) had their chronic immunosuppression regimen intensified. In 15 patients, a new drug or therapy was introduced (ECP in 6 patients, IVIg in 4 patients, mycophenolate in 3 patients and reinitiation of calcineurin inhibitor therapy in 2 patients). The remaining 10 patients had the dosing of existing therapies increased. Among the samples with borderline rejection, 18 (85.7%) patients had their immunosuppression increased, 2 (9.5%) received steroid pulse and in 1 case (4.8%) the patient was not weaned off prednisone. All the cases with changes to treatment had elevated dd-cfDNA levels.

**Figure 4:**
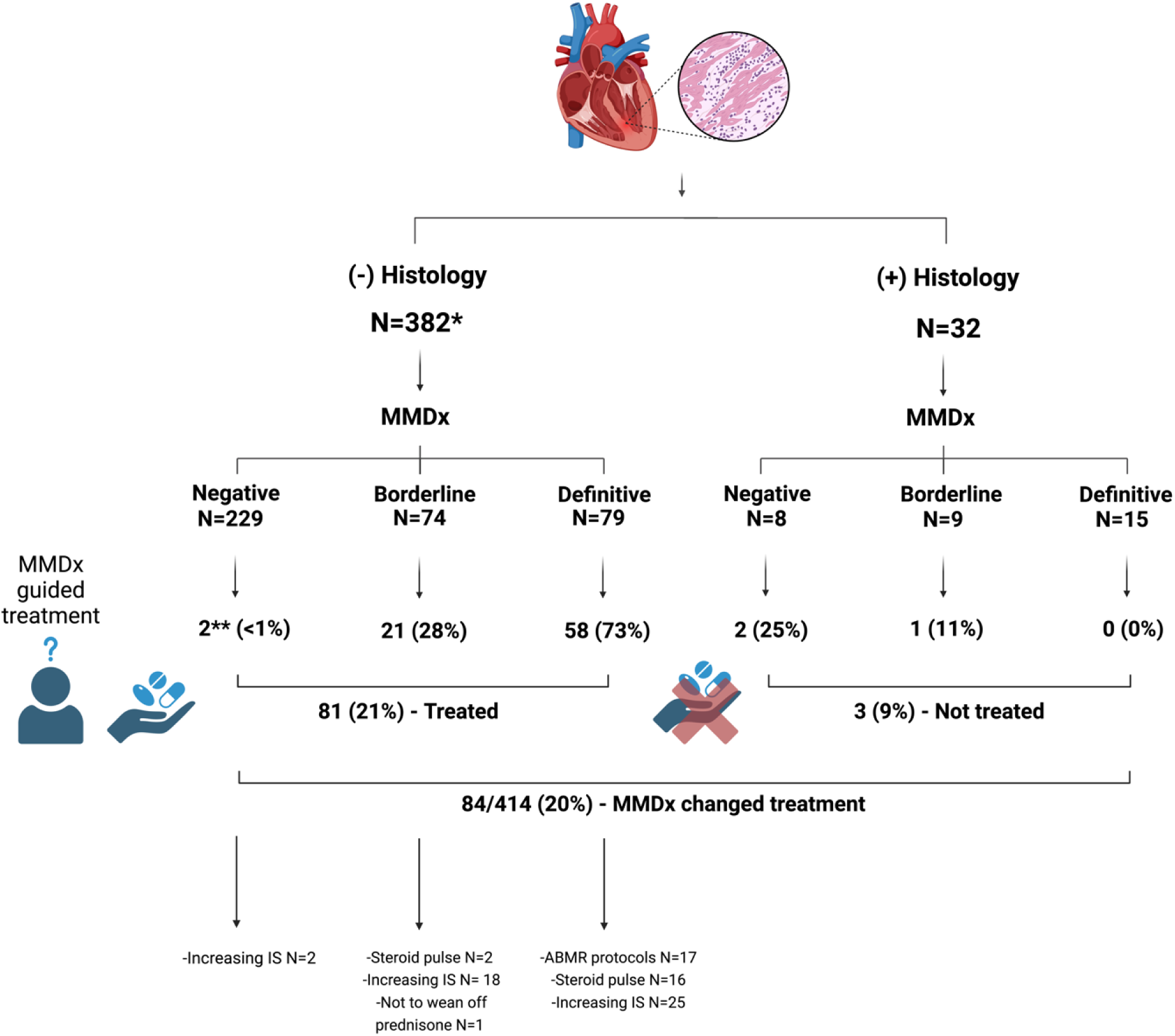
Histology and MMDx results. Treatment guided by MMDx results. IS: immunosuppression; *4 samples insufficient for ISHLT grading. **2 cases of parenchymal injury.

Conversely, in cases where histological rejection was present, the absence of rejection on MMDx also influenced clinical decisions. Specifically, in 3 out of 17 such cases, we either abstained from treatment or adopted a less aggressive approach based on MMDx results. This included 2 cases of pAMR 1i and 1 case of pAMR 2. **Figure 4**.

### Survival and graft function

At the 1-year follow-up, patients who had positive MMDx results and whose treatment was guided by those results (N=81) showed a survival rate of 87.0%, which was comparable to the survival rate of patients with negative MMDx results (89.0%). Conversely, among patients with positive MMDx results who did not undergo a change in therapy (N=98), the survival rate was significantly lower at 78.6% (log-rank p=0.02). **Figure 6**. No significant differences were observed in the occurrence of graft dysfunction between the groups (log-rank test; p = 0.66).

### Change in dd-cfDNA, DSA levels, and incidence of rejection during follow-up

An improvement in dd-cfDNA levels, defined as a 25% reduction following treatment, was observed in 76% of patients who were treated based on MMDx results. Conversely, only 36% of patients with positive MMDx who did not receive treatment showed a reduction in dd-cfDNA levels (p=0.04). Improvement in DSA was observed in 28% of cases in the active intervention group compared to 10% in the group without active treatment (p=0.012). The median time from MMDx to follow-up DSA was comparable between the groups: 29 days [19-58] in the intervention group vs 36 days [24-73.5] in the non-intervention group (p=0.126). The rates of rejection during follow-up were similar between the groups, both by histology (6.25% vs 6.45%; p=0.66) and by MMDx (33.3% vs 37.5%; p=0.91).

### Change in gene expression transcripts

32 episodes of rejection on MMDx with a follow-up MMDx were identified. Following treatment, cases of TCMR showed significant improvement in TCMR’s rejection scores, all TCMR-related transcripts (**Figure 5; Table 4S**), and injury-related transcripts (QCMAT and HT1) on the following MMDx sample. Similarly, in cases of ABMR, all transcript-related clusters associated with ABMR (**Figure 5, Table 5S**) decreased after treatment. **Figure 2S** represents the remaining transcript set.

**Figure 5.**
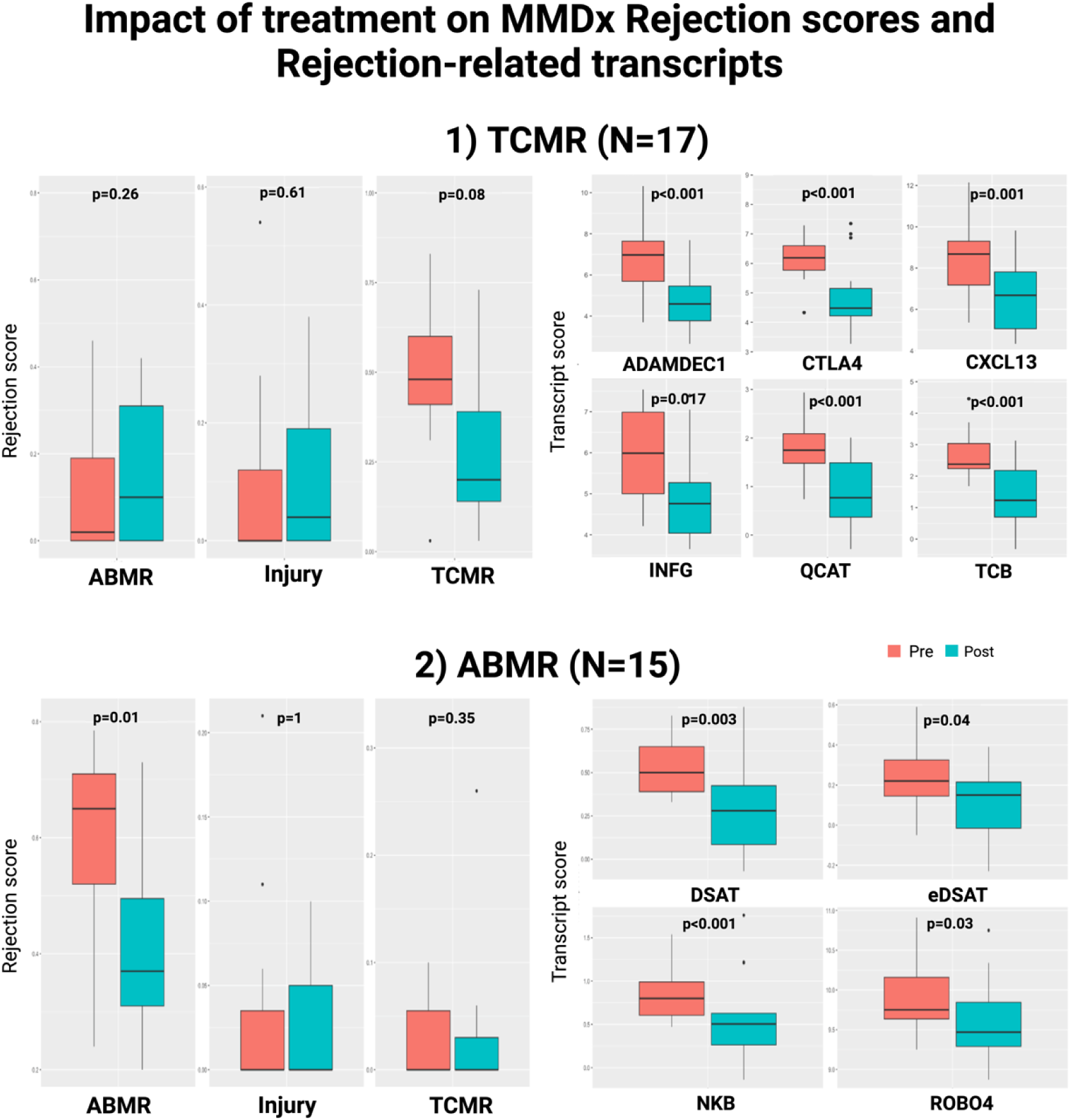
illustrates the changes in transcripts in the follow-up MMDx after treatment. The left panel represents overall rejection scores, including antibody-mediated rejection (ABMR), injury, and T-cell-mediated rejection (TCMR). The upper right panel shows a subset of transcripts related to TCMR (ADAMDEC1, CTLA4, CXCL12, INFG, QCAT, and TCB). The lower right panel displays a subset of transcripts related to ABMR (DSAT, eDSAT, NKB, ROBO4). All transcripts show improvement after treatment in their respective rejection groups.

**Figure 6:**
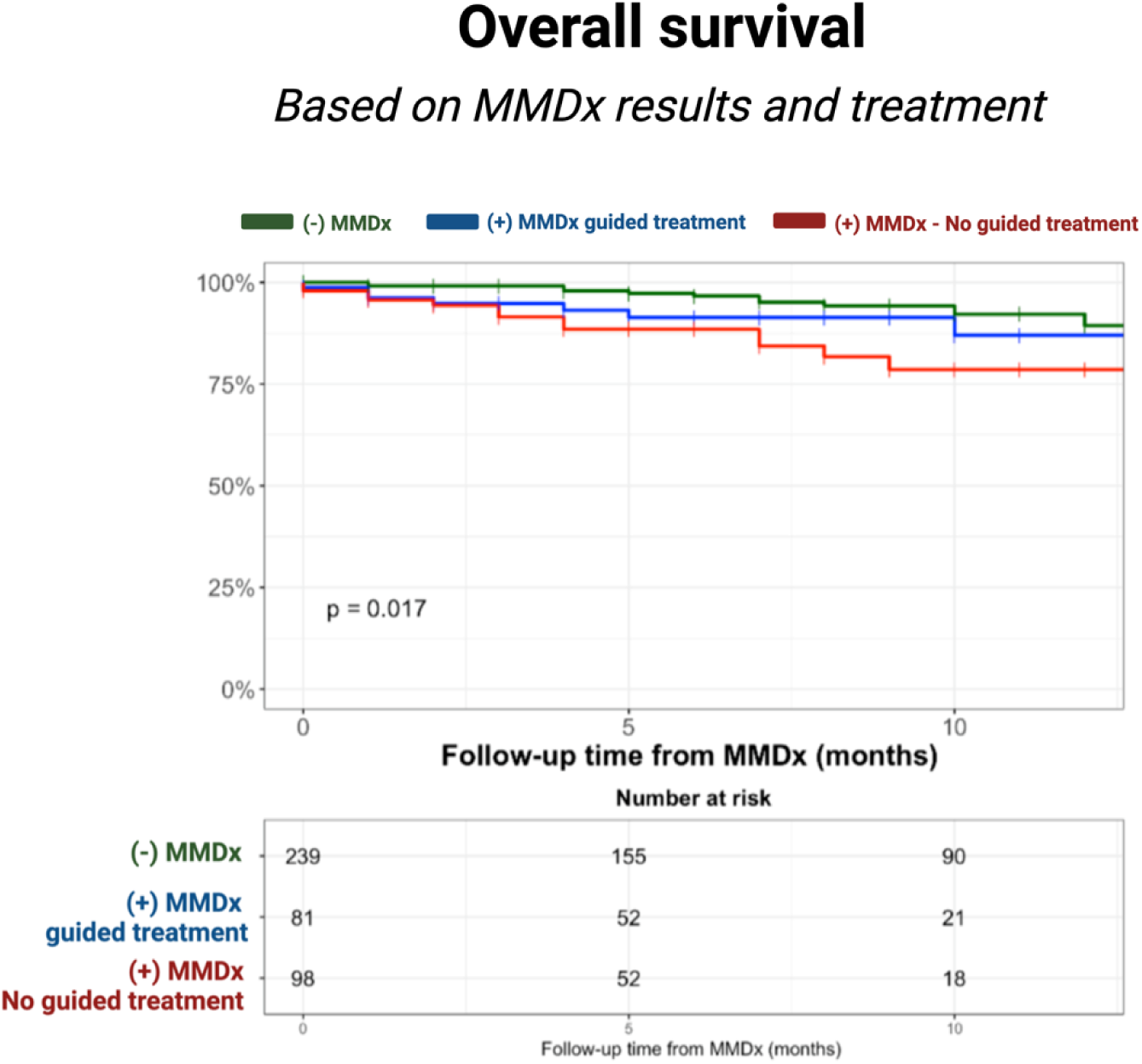
survival outcomes stratified based on MMDx results and MMDx-guided treatment.

### Kidney function

Median eGFR values at the time of biopsy for patients who received treatment based on MMDx were 72.5 mL/min/1.73m^2^ [50.3 – 90.0]. No significant differences were found in eGFR values at 1 month (75.7 [49.0 – 90.0]) or 3 months during follow-up (70.0 [51.3 – 90.0]; p=0.74). **Figure 3S.**

### Infection and malignancy

Among the 81 episodes of rejection treated based on MMDx results, 5 patients (6.2%) experienced a subsequent infection within 3 months requiring hospitalization. The infections included line-associated bacteremia related to ECP, infectious diarrhea, urinary tract infection, and soft-tissue cellulitis. Additionally, 7 patients (8.6%) experienced active viremia post-treatment, that did not require further intervention: 3 patients had BK viremia, 3 had EBV viremia, and 1 had CMV viremia, with a median viral load of 340 [130-385] copies/mL. One patient, who had initially been treated with pulse steroids due to TCMR was diagnosed with localized lung adenocarcinoma 13 months later.

## DISCUSSION

To our knowledge, this is the first evaluation of the clinical utility of MMDx in a cohort of heart transplant recipients with suspected rejection. Our major findings are as follows: 1) MMDx detected rejection three times more frequently than histology; 2) A pattern in dd-cfDNA levels was observed across cases with definitive, borderline, and negative MMDx results; the rates of rejection detected by MMDx were four times higher when both dd-cfDNA and pGEP levels were elevated; 3) Overall, immunosuppression was modified in response to MMDx in 20% of the cases and in 73% of the instances where histology was negative and MMDx showed rejection; 4) For patients with a positive MMDx, survival was superior in patients who underwent treatment guided by MMDx compared to those who did not receive treatment based on MMDx findings; 5) In the active treatment group, both dd-cfDNA levels, DSA and MMDx rejection-related transcripts showed significant improvement during follow-up.

While histological assessment has been pivotal in understanding rejection patterns and crucial for the evolution of transplantation, its limitations have become evident (5,7). The incorporation of new tools like dd-cfDNA and MMDx, combined with the high incidence of biopsy-negative rejection in clinical practice, has highlighted the need to reassess how we determine the presence of rejection in the contemporary era (6, 22–23). Several studies have shown that dd-cfDNA elevations precede histological rejection and that elevated dd-cfDNA levels correlates more strongly with molecular diagnosis than with conventional histology (9, 16, 18). This highlights the superior diagnostic precision of MMDx, especially in cases where there are discrepancies between histological and molecular findings, underscoring the enhanced accuracy of MMDx over histology (11, 22, 25). The recently published SHORE (Surveillance HeartCare Outcomes Registry) (24) highlighted the diagnostic utility of combining dd-cfDNA and pGEP in identifying ACR on histology. Notably, only 9.3% of samples with both elevated dd-cfDNA and pGEP showed histological evidence of rejection. Whether these low rates reflect a lack of specificity of dd-cfDNA or the low sensitivity of histology for detecting rejection remains a subject of debate. Our study found that the utilization of MMDx resulted in a rejection rate nearly three times higher than traditional histology (22.7% vs 7.7%). This significant increase suggests that MMDx may be more sensitive in detecting rejection than conventional histology. Moreover, samples with rejection identified by MMDx exhibited higher rates of positive dd-cfDNA results and elevated dd-cfDNA levels compared to those with negative MMDx results. This contrasts with histology, where dd-cfDNA levels did not significantly differ between rejection and no-rejection states. Interestingly, a trend in dd-cfDNA levels was observed across definitive rejection, borderline rejection, and negative MMDx cases, suggesting that rejection might be better understood as a continuous spectrum of myocardial injury rather than a binary condition. This is further supported by the observation that samples with rejection detected by both MMDx and histology exhibited the highest dd-cfDNA levels compared to samples where only one method detected rejection. This implies that MMDx may be more effective in capturing this continuum of myocardial injury than traditional histology, potentially providing a more nuanced and accurate assessment of rejection.

The similarity in the presence of DSA and higher MFI antibody titers between cases of definitive and borderline rejection, despite lower levels of dd-cfDNA and rejection rates on histology in borderline cases, suggests that borderline changes may represent an early stage of rejection rather than a benign finding (14,18). Indeed, in our cohort, cases of borderline rejection were followed by treatment in approximately 1 out of 3 cases. This continuum perspective aligns with findings from the Trifecta-Heart study (18), which reported higher dd-cfDNA levels in definitive rejection cases compared to those with borderline changes or negative MMDx. These results mirror previous observations from kidney biopsies (17,25), reinforcing the correlation between non-invasive markers and molecular diagnostics.

In our cohort, the presence of a combined (+)pGEP/(+)dd-cfDNA was associated with a 4-fold increase in the rates of rejection on MMDx compared to samples with both negative results (36.7% vs 8.3%). Likewise, a (+)pGEP/(+)dd-cfDNA was nearly three times more frequent in samples with borderline rejection than those with normal MMDx (28.0% vs 10.3%; p<0.001). These data suggest that the combination of both tests may provide an improved performance over each test alone, similar to what was previously described in the SHORE registry (24).

In cases where histology was negative and MMDx showed rejection, ABMR was detected nearly twice as frequently as TCMR. This finding aligns with the INTERHEART study, which demonstrated that many biopsies initially classified as negative or having low-grade rejection based on histology actually exhibited some ABMR changes (14).

In our cohort, in nearly 75% of cases where histology was negative and MMDx detected rejection, patients were subsequently treated based on MMDx results. 29% of those cases received ABMR specific protocols. The increasing detection of ABMR is of particular interest, given the well-known limitations of histology in detecting pAMR. Improving the detection of ABMR may provide a key to addressing incipient inflammation and injury, potentially ameliorating or averting the progression of cardiac allograft vasculopathy (CAV). Although our study was underpowered to evaluate the impact of interventions on CAV outcomes due to the reduced follow-up time, patients whose treatment was guided by positive MMDx results demonstrated improved 1-year survival rates. In these cases, treatment was followed by a significant reduction in dd-cfDNA levels, DSA strength and MMDx-rejection transcripts. Importantly, this treatment approach did not adversely impact kidney function, and the rates of clinically relevant infectious complications were acceptable.

Taken together, these findings highlight the potential role of MMDx in guiding therapeutic interventions and question whether histology should remain the gold standard for rejection diagnosis. Nevertheless, longer follow-up and carefully designed trials are necessary to corroborate our findings. Future research should focus on whether acting on early signs of rejection will play a crucial role in preserving graft function and preventing the initiation and/or progression of CAV.

This study has several limitations. First, this is a single-center analysis which may limit its external generalizability. Second, we did not have a single pathologist review all histology, but the pathologists were blinded to MMDx results and clinical data in all cases. Third, dd-cfDNA/pGEP levels were not sent concurrently with biopsy in most cases although we limited the separation between these labs and MMDx to 31 days. Lastly, the response to MMDx results was not protocolized and was left up to the discretion of individual clinicians.

## Conclusion

Our results highlight the potential of MMDx to provide complementary information aiding in the diagnosis of allograft rejection and guiding clinical management. These data raise concerns about whether histology should continue to be used as the gold standard for rejection surveillance.

## Data Availability

Data could be available upon reasonable request

## Author contributions

all authors contributed significantly to the production of this manuscript and all authors have offered final approval of the submitted version.

## Acknowledgements

Andrea Fernandez Valledor received grant support by the Alfonso Martin Escudero Foundation.

## Abbreviations

HT: heart transplantation
MMDx: Molecular Microscope Diagnostic System
EMB: endomyocardial biopsy
dd-cfDNA: donor-derived cell free DNA
pGEP: peripheral gene-expression profiling
DSA: donor specific antibodies
ABMR: antibody-mediated rejection on MMDx
TCMR: T-cell mediated rejection on MMDx
pAMR: pathological antibody-mediated rejection
ACR: acute cellular rejection (histology).

## Supplementary material

**Figure 1S:**
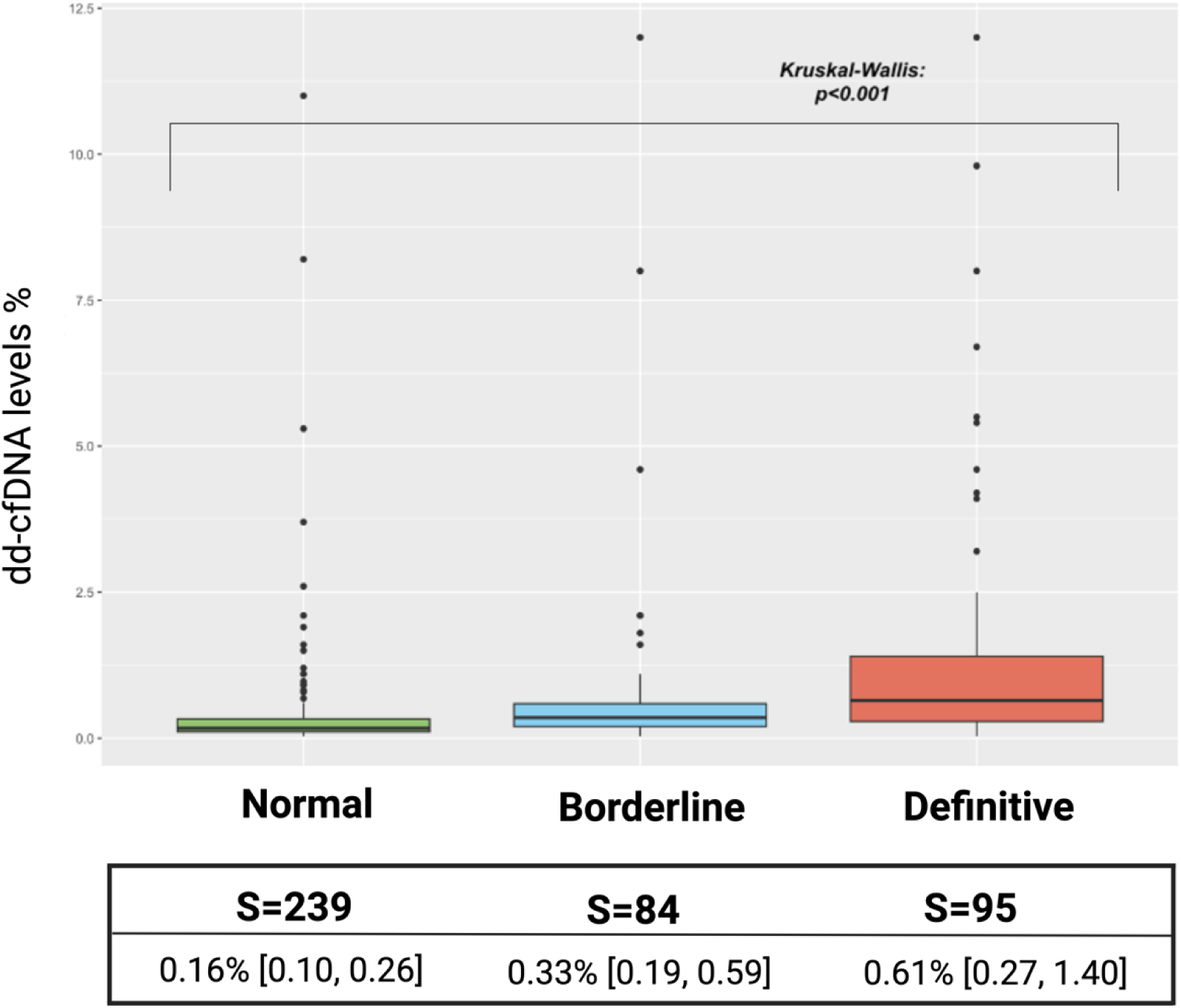
dd-cfDNA% levels according to MMDx results.

**Figure 2S:**
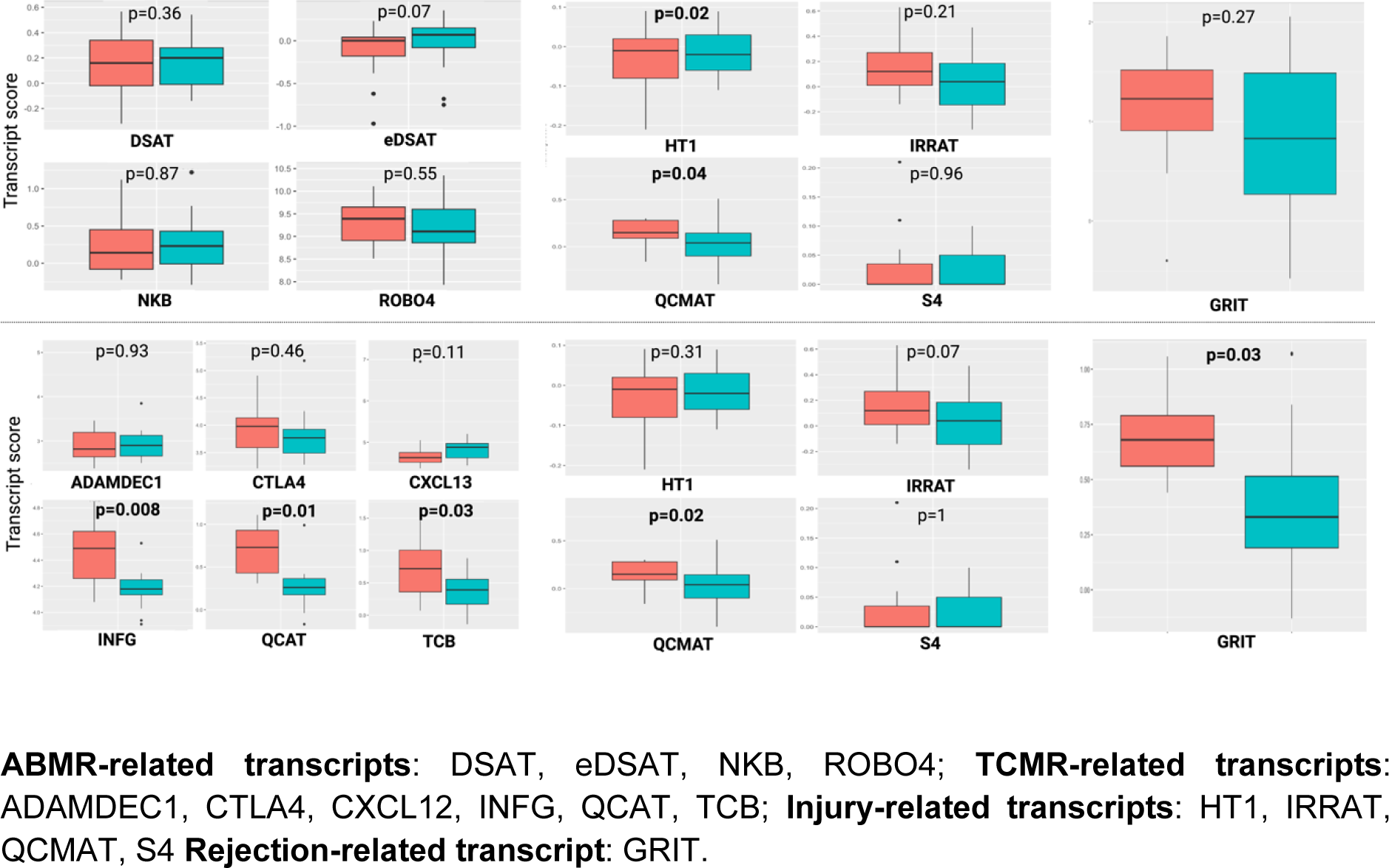
[Continuation of Figure 5]. Changes in Transcripts After Treatment; Upper group TCMR (N=17); Lower group: ABMR (N=15).

**Figure 3S:**
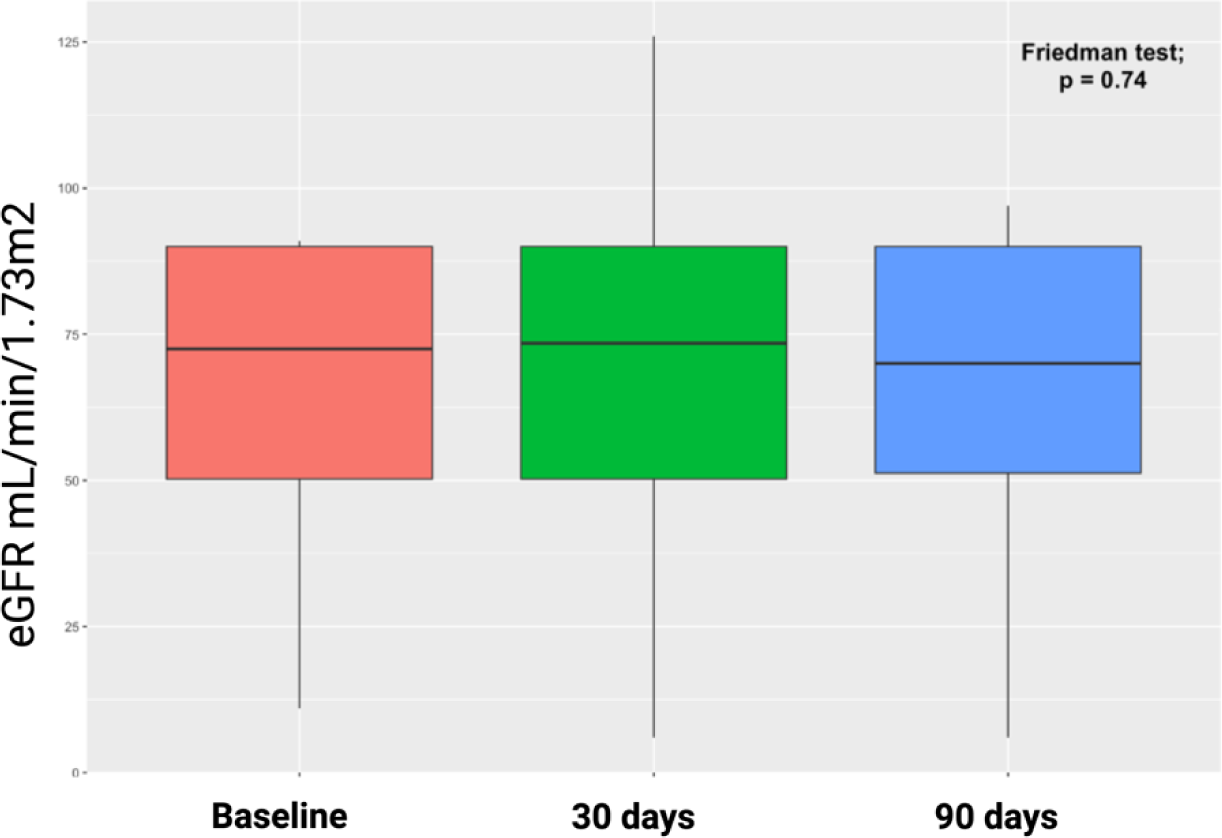
eGFR trends in the cohort of treated patients based on MMDx results at 1 month and 3 months after increasing immunosuppression.

**Table 1S:**
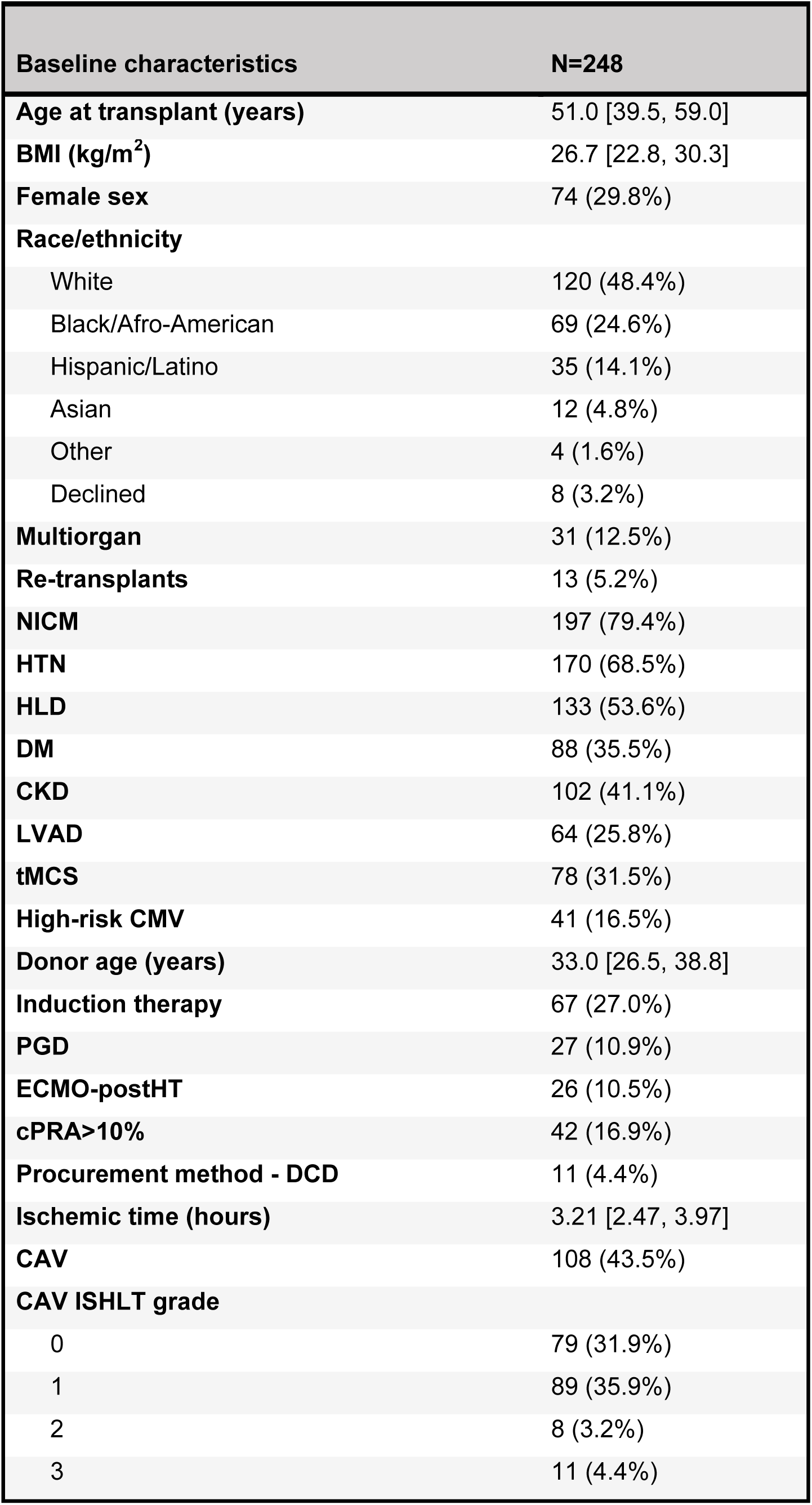
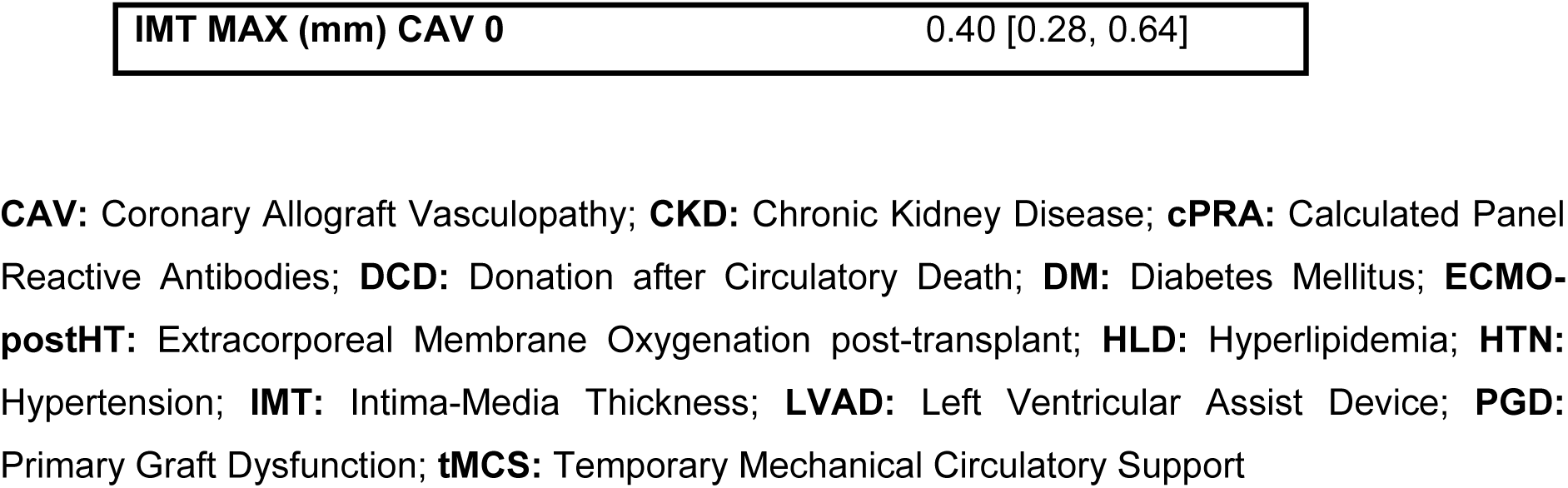
Baseline characteristics.

**Table 2S:** concordance and discordance between histology and MMDx.

|  | NR-MMDx<br>(-) Histology<br>(N=303) | NR-MMDx<br>(+) Histology<br>(N=17) | R-MMDx<br>(-) Histology<br>(N=79) | R-MMDx<br>(+) Histology<br>(N=15) | p-value |
| --- | --- | --- | --- | --- | --- |
| **dd-cfDNA≥ 0.12%** | 114 (66.7%) | 7 (70.0%) | 54 (91.5%) | 8 (88.9%) | **<0.001** |
| **dd-cfDNA≥ 0.20%** | 81 (47.4%) | 6 (60.0%) | 49 (83.0%) | 8 (88.9%) | **<0.001** |
| NA | 132 (43.6%) | 7 (41.3%) | 20 (25.3%) | 6 (40.0%) |  |
| **(+) pGEP** | 40 (13.2%) | 2 (11.8%) | 22 (27.8%) | 4 (26.7%) | **0.04** |
| NA | 151 (49.8%) | 9 (52.9%) | 27 (34.2%) | 7 (46.7%) |  |
| **dd-cfDNA %** | 0.19 [0.11, 0.37] | 0.28 [0.13, 0.43] | 0.59 [0.27, 1.30] | 0.78 [0.44, 4.60] | **<0.001** |
| **LVEF<50%** | 57 (18.8%) | 5 (29.4%) | 13 (16.5%) | 3 (20.0%) | 0.67 |
| **De-novo LVEF<50%** | 12 (3.9%) | 3 (17.6%) | 4 (5.1%) | 1 (6.7%) | 0.82 |
| **DSA** | 136 (44.9%) | 11 (64.7%) | 35 (44.3%) | 11 (73.3%) | **0.07** |
| DSA Class I | 51 (16.8%) | 9 (52.9%) | 16 (20.3%) | 6 (40.0%) | **<0.001** |
| DSA Class II | 122 (40.3%) | 8 (47.1%) | 28 (35.4%) | 10 (66.7%) | 0.15 |
| >4000 MFI | 67 (22.1%) | 9 (52.9%) | 16 (20.3%) | 10 (66.7%) | **<0.001** |
| **Histology results** |  |  |  |  |  |
| ACR>1R/1A | 0 (0%) | 6 (35.3%) | 0 (0%) | 4 (26.7%) |  |
| pAMR>0 | 0 (0%) | 11 (58.8%) | 0 (0%) | 11 (73.3%) |  |
| pAMR>1 | 0 (0%) | 7 (41.2%) | 0 (0%) | 4 (26.7%) |  |
| **MMDx results** |  |  |  |  |  |
| ABMR | 0 (0%) | 0 (0%) | 48 (60.8%) | 11 (73.3%) |  |
| TCMR | 0 (0%) | 0 (0%) | 26 (32.9%) | 4 (26.7%) |  |
| Mixed | 0 (0%) | 0 (0%) | 5 (6.3%) | 0 (0%) |  |
| Borderline | 73 (24.1%) | 9 (52.9%) | 0 (0%) | 0 (0%) |  |
| Injury | 25 (8.3%) | 3 (17.6%) | 0 (0%) | 0 (0%) |  |
| Normal | 205 (67.6%) | 5 (29.4%) | 0 (0%) | 0 (0%) |  |

\* **R:** definitive rejection on MMDx (TCMR, ABMR, Mixed Rejection); **NR:** no-definitive rejection on
MMDx (borderline, and negative MMDx).

**Table 3S:** sample characteristics stratified by MMDx results.

|  | 1 – Negative<br>(S=239) | 2 - Borderline<br>(S=84) | 3 – Definitive<br>(S=95) | p-value |
| --- | --- | --- | --- | --- |
| **ImmuKnow levels** | 338 [227, 512] | 288 [183, 446] | 283 [199, 430] | **0.01** |
| Time ImmuKnow to MMDx (d) | 7.50 [2.00, 15.0] | 8.00 [3.00, 15.0] | 8.50 [3.00, 14.0] | **0.002** |
| **eGFR mL/min/1.73m<sup>2</sup>** | 66.0 [44.0, 90.0] | 84.0 [56.0, 90.0] | 68.0 [41.0, 90.0] | **<0.001** |
| Time eGFR to MMDx (d) | 0 [0, 4.50] | 0 [0, 5.25] | 1.00 [0, 7.50] | **<0.001** |
| **NT-proBNP pg/dl** | 882 [341, 3100] | 628 [270, 2070] | 875 [384, 2630] | 0.40 |
| Time NT-proBNP to MMDx<br>(d) | 4 [1.00, 10.0] | 4 [0, 11.0] | 6.00 [1.00, 12.0] | 0.31 |
| **Hs-Troponin T (TnT) ng/l** | 38 [15.3, 81.5] | 32 [0.19, 57.5] | 17.0 [11.0, 35.0] | **<0.001** |
| Time hs-TnT to EMB (d) | 3.00 [1.00, 12.0] | 7.00 [0.50, 14.0] | 6.00 [1.00, 12.0] | 0.70 |
| **Abnormal RHC** | 64 (27.0%) | 19 (22.6%) | 33 (35.1%) | 0.34 |
| **Graft dysfunction** | 49 (20.5%) | 13 (15.5%) | 17 (17.9%) | 0.57 |
| **De-novo LVEF<50%** | 26 (10.2%) | 7 (8.3%) | 6 (6.3%) | 0.56 |
| **IS Treatment at MMDx** |  |  |  |  |
| Tacrolimus | 209 (87.4%) | 74 (88.1%) | 85 (89.5%) | **0.88** |
| Mycophenolate | 199 (83.3%) | 71 (84.5%) | 78 (82.1%) | 0.91 |
| Prednisone | 190 (79.5%) | 64 (76.2%) | 79 (83.2%) | 0.91 |
| mTOR inhibitors | 35 (14.6%) | 10 (11.9%) | 21 (22.1%) | 0.13 |
| Cyclosporin | 24 (10.0%) | 10 (11.9%) | 7 (7.4%) | 0.58 |
| Belatacept | 19 (7.9%) | 9 (10.7%) | 26 (27.4%) | **<0.001** |
| IVIG | 17 (7.1%) | 13 (15.5%) | 12 (12.6%) | 0.06 |
| ECP | 2 (0.8%) | 3 (3.6%) | 7 (7.4%) | **0.006** |

**D:** days; **ECP:** Extracorporeal Photopheresis; **eGFR:** estimated glomerular filtration rate; **hs-TnT:** Highly Sensitive Troponin T; **IS:** Immunosuppressive Treatment; **IVIG:** Intravenous Immunoglobulins; **mTOR inhibitors:** Inhibitors of Mammalian Target of Rapamycin; **RHC:** Right Heart Catheterization

**Table 4S:** Changes in transcripts in follow-up MMDx after treatment in the TCMR cohort.

|  | 1 - Before<br>(N=17) | 2 - After<br>(N=17) | p-value |
| --- | --- | --- | --- |
| **DSAT** | 0.160 [-0.0200, 0.340] | 0.220 [-0.0100, 0.280] | 0.36 |
| **eDSAST** | 0 [-0.180, 0.0400] | 0.0700 [-0.0800, 0.150] | 0.072 |
| **NKB** | 0.140 [-0.0800, 0.450] | 0.230 [-0.0100, 0.430] | 0.87 |
| **ROBO4** | 9.39 [8.91, 9.65] | 9.11 [8.86, 9.60] | 0.55 |
| **QCAT** | 1.61 [1.19, 2.00] | 0.770 [0.370, 1.70] | **<0.001** |
| **TCB** | 2.35 [2.14, 2.82] | 1.26 [0.750, 2.39] | **<0.001** |
| **ADAMDEC1** | 6.03 [5.49, 7.50] | 4.89 [4.01, 6.34] | **<0.001** |
| **CXCL13** | 8.68 [7.18, 9.12] | 7.01 [5.61, 7.81] | **0.0011** |
| **IFNG** | 5.93 [4.77, 6.43] | 4.82 [4.07, 5.27] | **0.017** |
| **CTLA4** | 5.96 [5.59, 6.49] | 4.69 [4.36, 5.88] | **<0.001** |
| **GRIT** | 1.23 [0.910, 1.52] | 0.830 [0.270, 1.49] | 0.27 |
| **HT1** | -0.200 [-0.270, -0.100] | -0.120 [-0.420, -0.0200] | **0.022** |
| **IRRAT** | 0.330 [0.200, 0.630] | 0.390 [0.0500, 0.600] | 0.21 |
| **S4** | 0 [0, 0.120] | 0.0400 [0, 0.190] | 0.96 |
| **QCMAT** | 0.640 [0.420, 0.840] | 0.380 [0.0700, 0.920] | **0.045** |
| **ABMR_SCORE** | 0.0200 [0, 0.190] | 0.100 [0, 0.310] | 0.26 |
| **TCMR_SCORE** | 0.480 [0.410, 0.600] | 0.200 [0.140, 0.390] | **0.008** |
| **INJURY_SCORE** | 0 [0, 0.120] | 0.0400 [0, 0.190] | 0.61 |

**ABMR-related transcripts:** DSAT, eDSAT, NKB, ROBO4; **TCMR-related transcripts:** ADAMDEC1, CTLA4, CXCL12, INFG, QCAT, TCB; **Injury-related transcripts:** HT1, IRRAT, QCMAT, S4 **Rejection-related transcript:** GRIT.

**Table 5S:**
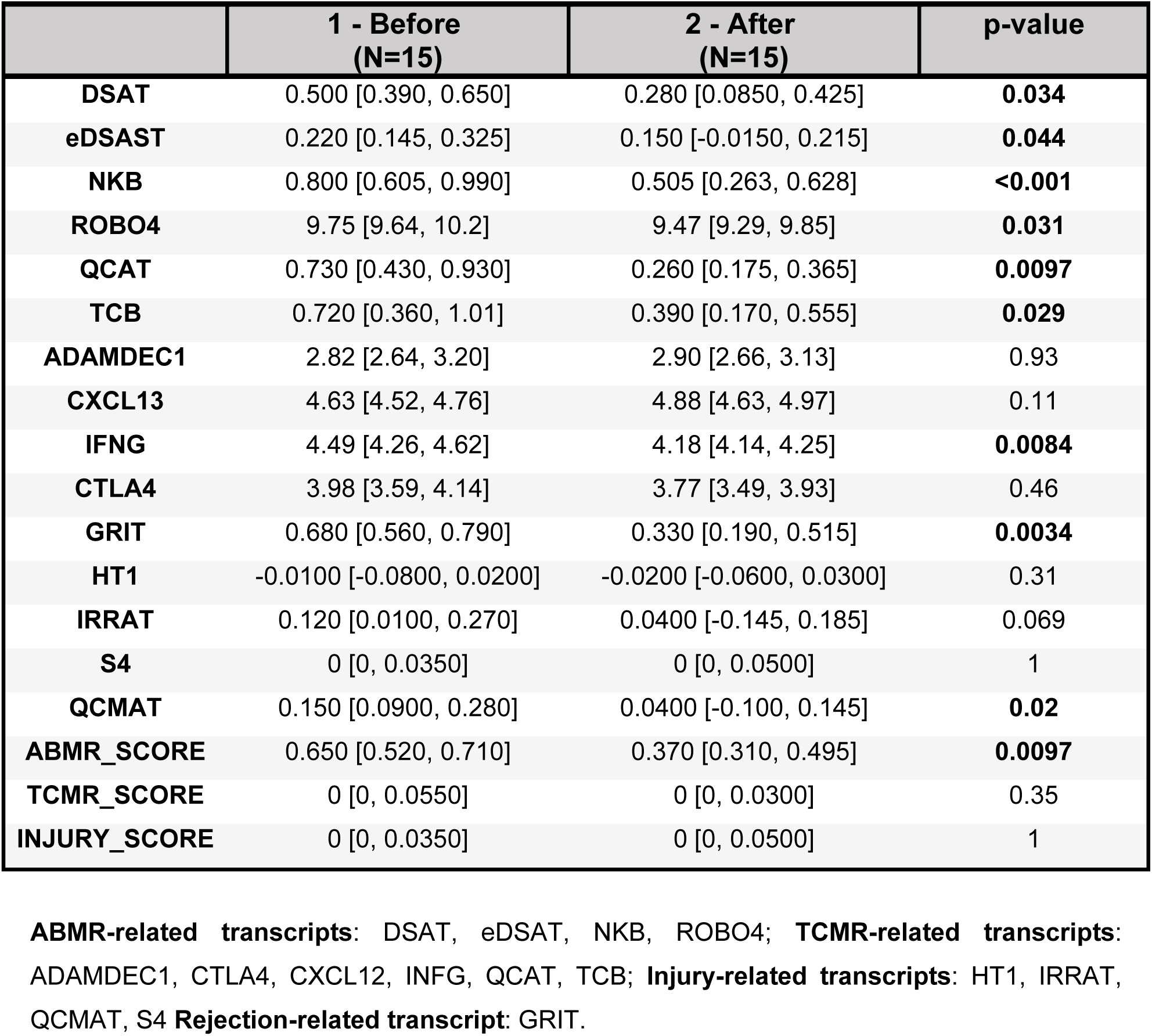
Changes in transcripts in follow-up MMDx after treatment in the ABMR group.

|  | 1 - Before<br>(N=15) | 2 - After<br>(N=15) | p-value |
| --- | --- | --- | --- |
| **DSAT** | 0.500 [0.390, 0.650] | 0.280 [0.0850, 0.425] | **0.034** |
| **eDSAST** | 0.220 [0.145, 0.325] | 0.150 [-0.0150, 0.215] | **0.044** |
| **NKB** | 0.800 [0.605, 0.990] | 0.505 [0.263, 0.628] | **<0.001** |
| **ROBO4** | 9.75 [9.64, 10.2] | 9.47 [9.29, 9.85] | **0.031** |
| **QCAT** | 0.730 [0.430, 0.930] | 0.260 [0.175, 0.365] | **0.0097** |
| **TCB** | 0.720 [0.360, 1.01] | 0.390 [0.170, 0.555] | **0.029** |
| **ADAMDEC1** | 2.82 [2.64, 3.20] | 2.90 [2.66, 3.13] | 0.93 |
| **CXCL13** | 4.63 [4.52, 4.76] | 4.88 [4.63, 4.97] | 0.11 |
| **IFNG** | 4.49 [4.26, 4.62] | 4.18 [4.14, 4.25] | **0.0084** |
| **CTLA4** | 3.98 [3.59, 4.14] | 3.77 [3.49, 3.93] | 0.46 |
| **GRIT** | 0.680 [0.560, 0.790] | 0.330 [0.190, 0.515] | **0.0034** |
| **HT1** | -0.0100 [-0.0800, 0.0200] | -0.0200 [-0.0600, 0.0300] | 0.31 |
| **IRRAT** | 0.120 [0.0100, 0.270] | 0.0400 [-0.145, 0.185] | 0.069 |
| **S4** | 0 [0, 0.0350] | 0 [0, 0.0500] | 1 |
| **QCMAT** | 0.150 [0.0900, 0.280] | 0.0400 [-0.100, 0.145] | **0.02** |
| **ABMR_SCORE** | 0.650 [0.520, 0.710] | 0.370 [0.310, 0.495] | **0.0097** |
| **TCMR_SCORE** | 0 [0, 0.0550] | 0 [0, 0.0300] | 0.35 |
| **INJURY_SCORE** | 0 [0, 0.0350] | 0 [0, 0.0500] | 1 |

**ABMR-related transcripts:** DSAT, eDSAT, NKB, ROBO4; **TCMR-related transcripts:** ADAMDEC1, CTLA4, CXCL12, IFNG, QCAT, TCB; **Injury-related transcripts:** HT1, IRRAT, QCMAT, S4 **Rejection-related transcript:** GRIT.

